# SARS-CoV-2 antibody signatures robustly predict diverse antiviral functions relevant for convalescent plasma therapy

**DOI:** 10.1101/2020.09.16.20196154

**Authors:** Harini Natarajan, Andrew R. Crowley, Savannah E. Butler, Shiwei Xu, Joshua A. Weiner, Evan M. Bloch, Kirsten Littlefield, Wendy Wieland-Alter, Ruth I. Connor, Peter F. Wright, Sarah E. Benner, Tania S. Bonny, Oliver Laeyendecker, David Sullivan, Shmuel Shoham, Thomas C. Quinn, H. Benjamin Larman, Arturo Casadevall, Andrew Pekosz, Andrew D. Redd, Aaron A.R. Tobian, Margaret E. Ackerman

**Affiliations:** Department of Microbiology and Immunology, Geisel School of Medicine at Dartmouth, Dartmouth College, Hanover, NH, USA; Thayer School of Engineering, Dartmouth College, Hanover, NH, USA; Department of Pathology, Johns Hopkins School of Medicine, Baltimore, MD, USA; W. Harry Feinstone Department of Molecular Microbiology and Immunology, Johns Hopkins Bloomberg School of Public Health, Baltimore, MD, USA; Department of Pediatrics, Geisel School of Medicine at Dartmouth, Dartmouth-Hitchcock Medical Center, Lebanon, NH, USA; Department of Epidemiology, Johns Hopkins Bloomberg School of Public Health, Baltimore, MD, USA; Department of Medicine, Division of Infectious Diseases, Johns Hopkins School of Medicine, Baltimore, MD, USA; Division of Intramural Research, National Institute of Allergy and Infectious Diseases, National Institutes of Health, Bethesda, MD, USA

**Author notes:** Contributed equally (Harini Natarajan, Andrew R. Crowley, Savannah E. Butler). Co-senior authors (Andrew D. Redd, Aaron A.R. Tobian, Margaret E. Ackerman). Corresponding Author Margaret E. Ackerman, 14 Engineering Drive, Hanover, NH 03755, (ph).

**Keywords:** Convalescent plasma, SARS-CoV-2, COVID-19, neutralization, functional antibody response, ADCC, phagocytosis

## Abstract

Convalescent plasma has emerged as a promising COVID-19 treatment. However, the humoral factors that contribute to efficacy are poorly understood. This study functionally and phenotypically profiled plasma from eligible convalescent donors. In addition to viral neutralization, convalescent plasma contained antibodies capable of mediating such Fc-dependent functions as complement activation, phagocytosis and antibody-dependent cellular cytotoxicity against SARS-CoV-2. These activities expand the antiviral functions associated with convalescent plasma and together with neutralization efficacy, could be accurately and robustly from antibody phenotypes. These results suggest that high-throughput profiling could be used to screen donors and plasma may provide benefits beyond neutralization.

---

Since its emergence in 2019, SARS-CoV-2 has spread rapidly and infected over 25 million individuals worldwide. As the medical community has mobilized to identify effective therapies to combat the virus, treatment with convalescent plasma derived from individuals who have recovered from COVID-19 has emerged as a potential therapeutic intervention^1,2^. Preliminary evidence suggests that patients treated early with convalescent plasma show improved survival and reduced viral load^1,3–5^. Case reports also suggest that convalescent plasma may be an effective antiviral in patients with impaired immunity^6,7^. However, antibody responses resulting from infection are highly variable in magnitude and character^8–10^. While the Expanded Access Program demonstrated efficacy of convalescent plasma in a dose-response effect of neutralizing titers, units with low titers were also efficacious suggesting that there are other contributing activities that have not been measured^11^. Thus, a better understanding the breadth and spectrum of antiviral activities of the humoral immune response is critical to maximizing the success of this promising intervention.

Beyond antibody titer, neutralizing antibody responses, which are typically elevated in association with severe disease, have exhibited wide variation among individuals^8,12^. Because antibodies directed against the receptor-binding domain of the fusogenic spike (S) protein can block the interaction of the spike with the angiotensin converting enzyme 2 (ACE2) receptor of airway epithelial cells and have demonstrated the ability to inhibit infection *in vitro*^13,14^ and *in vivo*^15–17^, these responses have been a key target in development of vaccines to prevent SARS-CoV-2 and monoclonal antibodies to treat COVID-19 disease^12^. Recent data suggest the frequency of neutralizing antibodies (nAbs) within the total humoral response could be quite low^18,19^, and that many antibodies are directed toward non-neutralizing epitopes within more conserved regions of the S protein^20,21^. One possible explanation for this observation is that SARS-CoV-2 may reactivate a pre-existing B cell population generated from prior exposures to endemic coronaviruses (CoV). Consistent with this hypothesis, some individuals who recovered from infection by SARS-CoV-1 exhibited elevated antibody titers against common coronaviruses^22,23^, particularly against OC43^24^, which falls within the betacoronavirus genus together with SARS-CoV-1 and CoV-2. This rapid recall response is suggestive of “original antigenic sin”, a phenomenon widely reported in influenza in which B cells targeting conserved but typically not cross-neutralizing epitopes are reactivated with the effect of reducing the generation of novel antibodies against neutralizing epitopes on the new virus^25,26^. Collectively, the known diversity of responses among convalescent individuals, as well as the possible influence of prior endemic CoV infection on induction of potently neutralizing antibodies to SARS-CoV-2, further motivate exploration of the range of responses in potential plasma donors.

In the absence of sufficient levels of direct antiviral activity via Ab-mediated blocking, the burden for humoral protection falls to the extra-neutralizing effector functions, which are initiated by the relatively constant domain (Fc) of virus-specific antibodies and executed by innate immune cells and the complement cascade. By engaging soluble and cell surface-expressed Fc receptors, antibodies can trigger a variety of functions such as phagocytosis, cellular cytotoxicity, and complement deposition, which play an important role in clearing diverse viral infections^27^. In the context of SARS-CoV-1, antibody-mediated phagocytosis has been observed to play a critical role in clearing infection *in vivo*, even in the context of exisiting potent neutralization activity^28^. Such findings indicate that understanding the ability of convalescent donor plasmas to elicit these effector functions in diverse infected subjects may be key to successful treatment.

This study evaluated the biophysical and functional evaluation of plasma samples from 126 eligible convalescent donors to define the features of functionally potent plasma antibody responses of high relevance to convalescent plasma donor selection, and relevant to the potential resistance of convalescent individuals to reinfection.

## Results

### Biophysical Characterization of SARS-CoV-2 Convalescent Plasma

Convalescent plasma samples from 126 eligible donors from the Baltimore/Washington D.C. area (Johns Hopkins Medical Institutions, JHMI cohort)^8^ and serum samples from 15 naïve controls and 20 convalescent subjects from New Hampshire (Dartmouth-Hitchcock Medical Center, DHMC cohort)^29^ serving as a validation cohort were collected (**Supplemental Table 1**). Antibody responses were evaluated using an Fc array assay that assesses both variable fragment (Fv) and Fc domain characteristics of antibodies^30^, which was customized to assess responses across a panel of SARS-CoV-2 and other endemic and pathogenic CoV antigens. This panel consisted of the nucleocapsid (N) protein, stabilized trimeric spike protein (S-2P)^31^, spike subdomains including S1 and S2, the receptor binding domain (RBD), and the fusion peptide from SARS-CoV-2; spike and S1 proteins from CoV associated with other epidemics (SARS-CoV-1, MERS); widely circulating CoV (OC43, HKU1, 229E, NL63); the closely related bat CoV, WIV1; and influenza HA and herpes simplex virus gE as controls. Characterization extended beyond antigen specificity to include antibody isotype, subclass, and propensity to bind Fc receptors (FcRs).

Diverse SARS-CoV-2-specific immunoglobulin isotypes and subclasses, particularly IgG1 and IgG3, IgA, and IgM, were elevated in SARS-CoV-2 convalescent subjects across different epitope and antigen specificities (**Figure 1A, Supplemental Figures 1-3**). Robust responses to stabilized spike (S-2P) and N were apparent, and lower magnitudes of responses were detected to functionally relevant RBD and fusion peptide domains. Relative to naïve subjects, the levels of OC43 S protein-specific IgG and IgA responses were elevated among convalescent donors (**Figure 1B, C**), suggesting the possibility that pre-existing cross-reactive antibodies may have been boosted by SARS-CoV-2 infection. Responses to other endemic CoV were generally more comparable between convalescent and naïve subjects. However, elevated IgG1 responses to the spike protein of other endemic CoV, including 229E, HKU1, and NL63 were observed.

**Figure 1.**
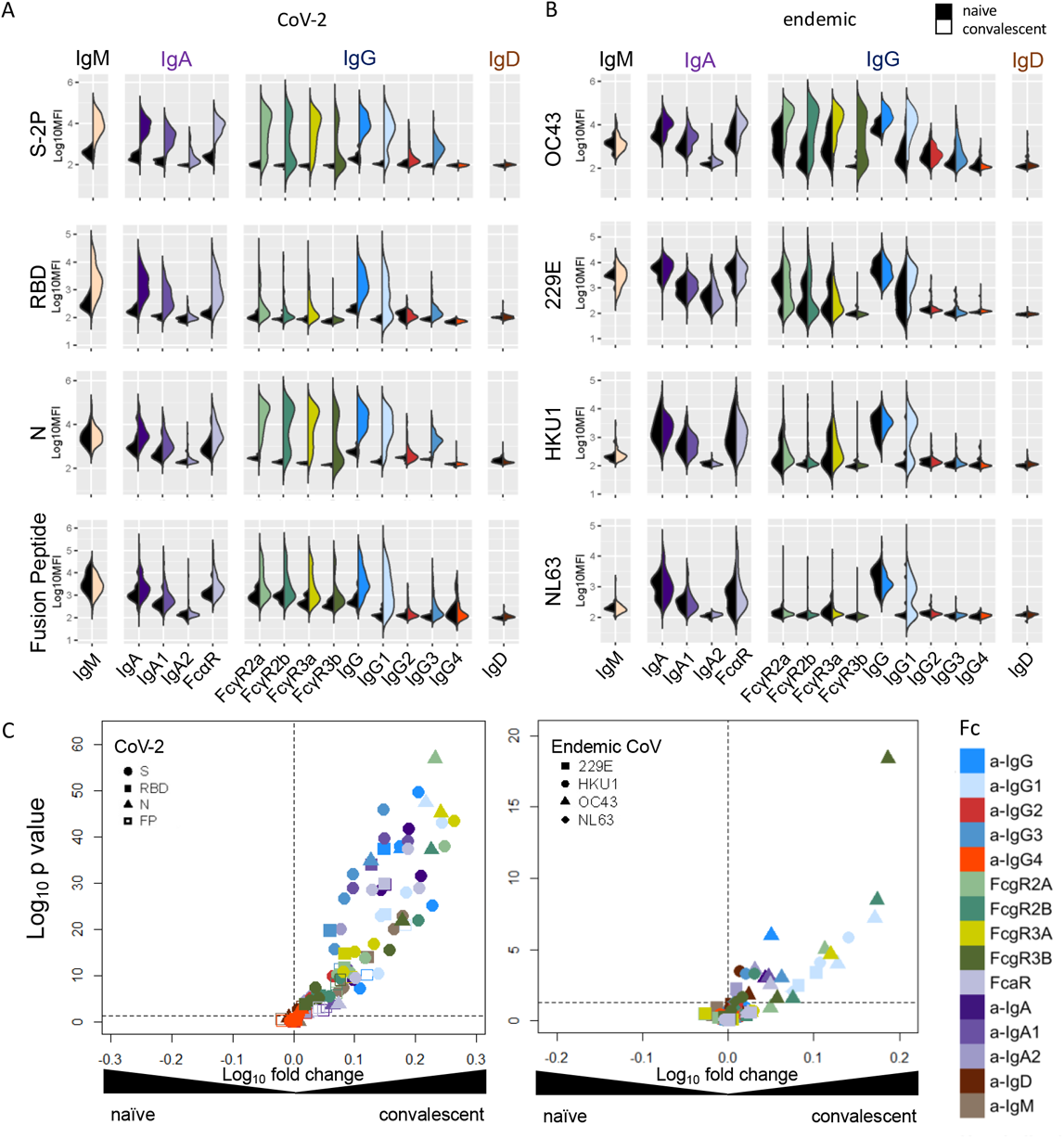
Antibody Responses in Convalescent Plasma. **A-B**. Fc array characterization of antibodies to SARS CoV-2 antigens (**A**) and endemic CoV (**B**) across antibody subclasses, isotypes, and binding to FcR in naïve (serum) and convalescent (plasma) donors. **C**. Volcano plot of fold change and significance of differences between convalescent and naïve subject antibody response features specific for SARS-CoV-2 (left) and endemic CoV (right).

To understand how the different facets of the Ab response related to one another, hierarchical clustering was performed on the biophysical antibody profiles of convalescent subjects who were hospitalized or not hospitalized, and naïve subjects. Subjects showed extensive variability in the SARS-CoV-2-specific Ab response magnitude and character (**Figure 2**). High levels of IgG were observed in many individuals, although a small number of convalescent donors appeared not to seroconvert despite being PCR+ on initial diagnosis and sample collection conducted an average of 43 days subsequently. Similarly, there was variability in the IgA responses raised in SARS-CoV-2-convalescent subjects, with some individuals showing relative higher IgA responses and others biased toward relatively higher IgG. Distinctions in antibody responses between subjects were apparent among antigen specificities, with some subjects mounting robust IgA responses to N or fusion peptide, but many more favoring the various S domains. Responses among hospitalized donors tended to be elevated in SARS-CoV-2-specific IgG as compared to subjects with less severe disease. Variable responses to endemic CoVs were observed in both the naïve and convalescent plasma. IgM and IgG1 responses were clustered across diverse endemic CoVs, whereas other features were grouped primarily by CoV strain across a wider diversity of Fc characteristics.

**Figure 2.**
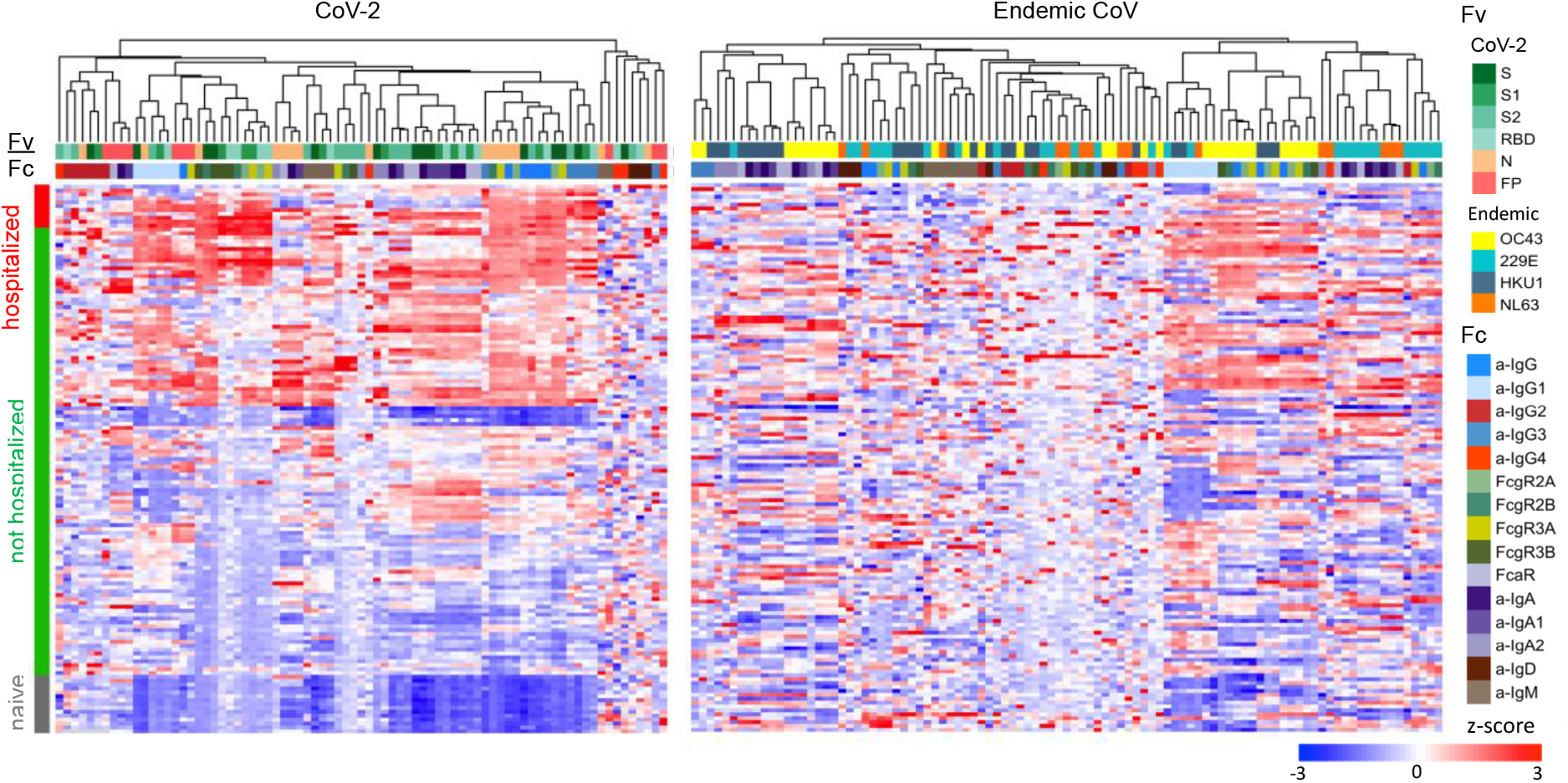
Humoral response profiles. Heatmap of filtered and hierarchically clustered SARS-CoV-2-specific antibody response features that were significantly elevated among convalescent donors (left) and endemic CoV-specific antibody features that were elevated above background (right). Responses were scaled and centered per feature and truncated at +/- 3 SD. Antigen specificity (Fv), Fc characteristics (Fc), and subject group are indicated in the color bars.

Consistencies apparent in the subjects with high IgG responses specific to SARS-CoV-2 and those with high responses toward OC43 suggested the value of a broader investigation of correlative relationships between responses to distinct CoV strains (**Figure 3A**). SARS-CoV-2- and OC43-specific IgG responses were positively correlated, consistent with cross-reactivity and boosting of a recall response. Whereas SARS-CoV-2-specific IgG and IgA responses were observed to be negatively correlated in a prior study^29^, evidence of an either/or aspect between isotypes was not observed. Strong and significant positive correlations were generally not observed between these isotypes in this cohort. To further understand the relationship between the SARS-CoV-2 antibody response and the elevation of responses toward some endemic CoVs, we compared OC43, HKU1, NL63, and 229E-specific IgG and IgA responses. We examined IgG responses to two different variants of the OC43 CoV spike protein, the wild-type spike protein (S) and a stabilized form thought to remain in a more native conformation (S-2P)^32^. OC43 S, but not S-2P-specific IgG responses were elevated in both convalescent donor cohorts as compared to naïve subjects (**Figure 3B**). Correlations between SARS-CoV-2-specific IgG and IgA and OC43-specific IgG and IgA were measured; compared to OC43 S-2P, responses to OC43 S were better correlated to SARS-CoV-2-specific antibody responses (**Figure 3C-D**). The increased correlation of SARS-CoV-2-specific responses to wild-type OC43 S relative to OC43 S-2P suggests that these boosted and/or cross-reactive antibodies may target primarily non-neutralizing epitopes that are presented by S, but not available on the stabilized S-2P trimer. Consistent with this hypothesis, OC43 S-specific responses were better correlated with responses to SARS-CoV-2 S2, which exhibits greater homology to endemic CoVs, than to SARS-CoV-2 RBD, which is recognized by many neutralizing antibodies.

**Figure 3.**
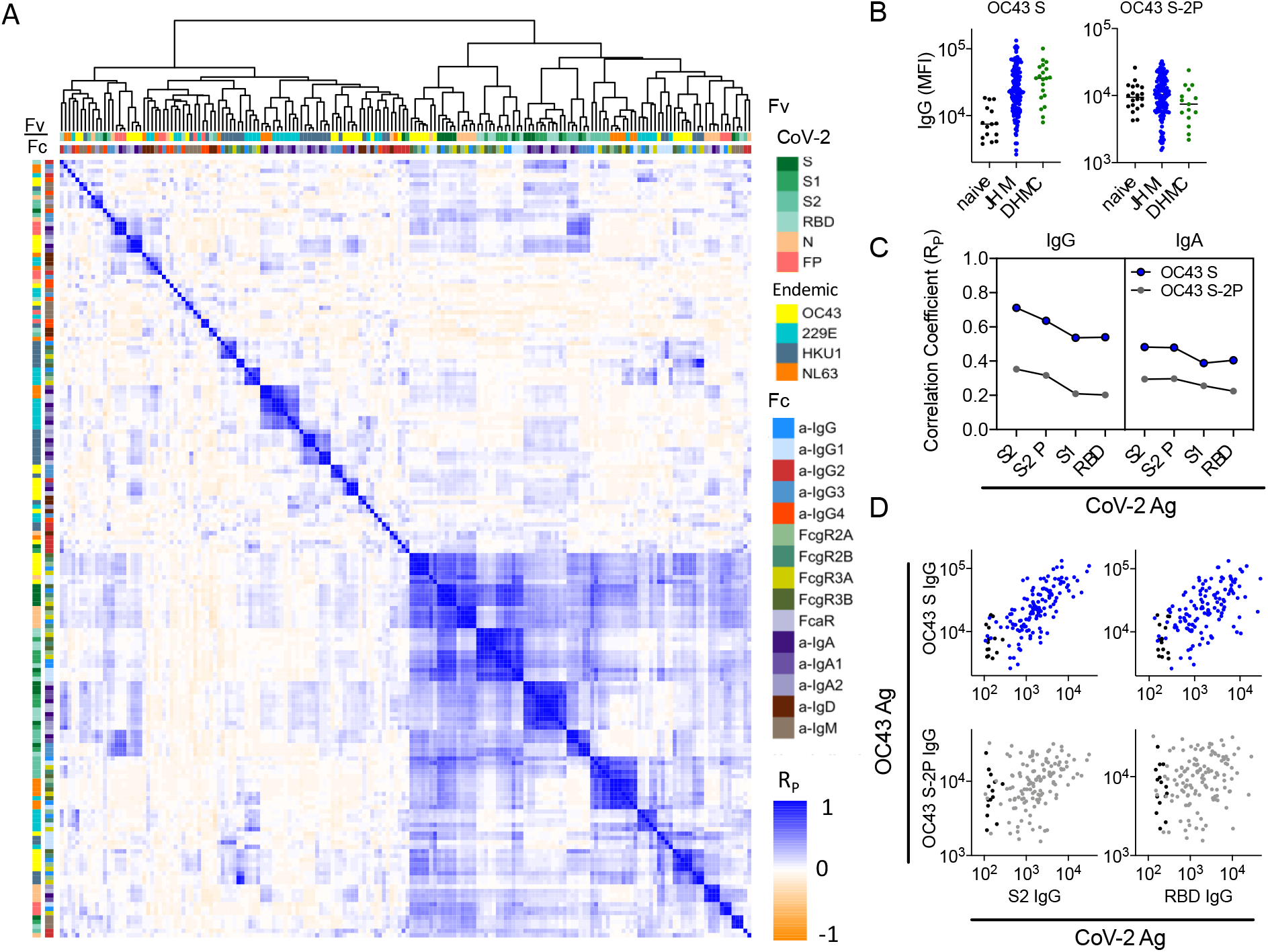
Correlative relationships between antibody features in convalescent plasma. **A**. Correlation matrix of relationships between filtered Ab features. Antigen specificity (Fv) and Fc characteristics (Fc) are indicated by the color bars. Filtered features are hierarchically clustered and Pearson coefficients (Rp) are shown. **B**. Comparison between IgG levels in naïve, DHMC, and JHMI cohort samples to OC43 S and OC43 S-2P. **C**. Correlation (Rp) between IgG and IgA specific to SARS-CoV-2 antigens and IgG and IgA specific to OC43 S and OC43 S-2P. **D**. Scatterplots of IgG responses specific to OC43 S and OC43 S-2P versus CoV-2 S2 and RBD. Naïve subjects are shown in black and convalescent donors are shown in blue (OC43 S) and gray (OC43 S-2P).

Broadening this analysis to more distantly-related CoV, relative to HKU1 and 229E S1-specific antibodies, S-2P-specific antibodies were correlated more strongly to responses to SARS-CoV-2 antigens (**Supplemental Figure 4**). Again, responses to endemic CoV were generally better correlated to those targeting the S2 rather than RBD domain of SARS-CoV-2, consistent with domain homology. These relationships further suggest that potentially cross-reactive antibodies may be more likely to recognize the S2 domain, less likely to recognize S1 or RBD domain, and therefore unlikely to be neutralizing.

### Relationship Between Antibody Characteristics and Clinical Characteristics

To determine how humoral immune responses related to clinical characteristics, differences in the antibody response toward SARS-CoV-2 and endemic CoV associated with sex, age, and hospitalization status were evaluated (**Supplemental Figure 5**). While distinctions in responses toward endemic CoV based on clinical characteristics were infrequently observed and relatively weak, SARS-CoV-2 IgG, IgA, and Fc*γ*R-binding antibodies were significantly elevated in older and male subjects, characteristics which are considered risk factors for more severe disease. Confounding effects associated with covariation in clinical characteristics were not observed, suggesting the independence of these subject characteristics. Elevated SARS-CoV-2-specific IgG and Fc*γ*R-binding antibody features were also observed in hospitalized subjects, consistent with prior studies^8,29,33^, and the possibility that IgG responses may drive disease enhancement^26,34,35^. However, despite being associated with both age and sex risk factors, elevated IgA features were not observed in hospitalized subjects, consistent with the possibility that IgA responses may contribute to milder infection^29^.

### Distinctinctions between subjects defined by humoral response profiles

To define similarities and differences among donors more globally, dimensionality reduction was performed on biophysical features using Uniform Manifold Approximation and Projection (UMAP)^36^. Subjects were distributed across the antibody biophysical profile landscape into a set of four distinct clusters (**Figure 4A**). Though hospitalized subjects were observed in multiple clusters, they were most prevalent in cluster 4 and adjacent regions of clusters 2 and 3. To understand aspects of the humoral response that distinguished each cluster, univariate testing was performed to determine and depict which Fc array features were distinct for individual clusters (**Figure 4B**). Relative responses for these features among convalescent donors in each group reflect differences in the magnitude of the response, with cluster 1 having lower humoral responses to SARS-CoV-2 antigens in general, clusters 2 and 3 exhibiting intermediate responses, and cluster 4 typically showing globally elevated antibody responses. Clusters 2 and 3, which both presented with intermediate response magnitudes, were distinguished by relative differences in IgG1 versus IgA responses. Reduced IgG1 responses in cluster 2 were not unique to SARS-CoV-2, but extended across diverse endemic CoV spike proteins (**Figure 4C**).

**Figure 4.**
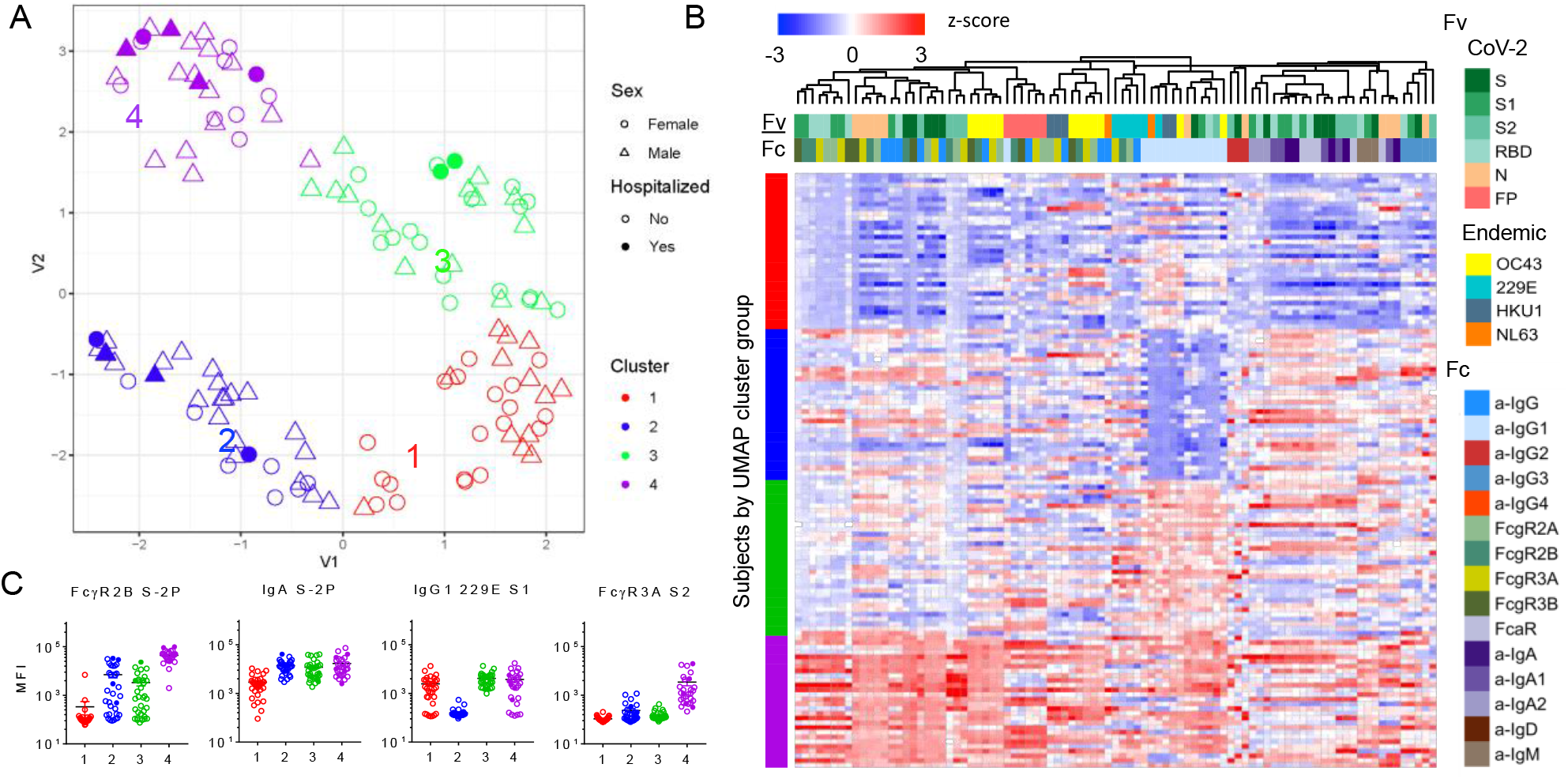
Distinctions among convalescent plasma donors in humoral response profile. **A**. UMAP analysis of subjects. Position in variable (V1, V2) space indicate similarity or distinctions in antibody response. Symbols and color indicate subject sex, hospitalization status, and cluster. **B**. Heatmap of distinct features by group. Antigen specificity (Fv) and Fc characteristics (Fc) are indicated by the color bars. **C**. Exemplary boxplots of distinct features.

### Antibody effector function and feature correlations

To explore the biological functions of antibodies in convalescent donors, both neutralizing and extra-neutralizing activities were evaluated and reported across clustered subject groups (**Figure 5A**). Consistent with the overall SARS-CoV-2 rank order of antibody response magnitude, neutralization activity against live virus was highest among cluster 4 and lowest among cluster 1 subjects. While antibody-dependent cell-mediated phagocytosis (ADCP), Fc*γ*RIIIa-activation as a surrogate for antibody-dependent cellular cytotoxicity (ADCC), and antibody-dependent complement deposition (ADCD) elicited by RBD-specific antibodies were highest among cluster 4 and lowest among cluster 1, correlative relationships between functions showed distinctions among these antiviral activities (**Figure 5B**). ADCP, known to play an important role in viral clearance in a SARS-CoV-1 disease mouse model^28^, and ADCC were highly correlated with each other (R^P^=0.82), and moderately correlated with neutralization (R^P^=0.64 and 0.57, respectively). Complement activation, which has been associated with increased inflammation and disease pathology in a mouse model of SARS-CoV-1^37^, as may also be the case in COVID-19 disease^38,39^, was less well correlated with other activities. Antibody-mediated deposition of complement component C3b on SARS-CoV-2 S1 and RBD showed greater distinctions among clusters than did deposition on trimeric S-2P, whose more uniform activity profile across groups is consistent with a greater contribution of recalled responses against endemic CoV.

**Figure 5.**
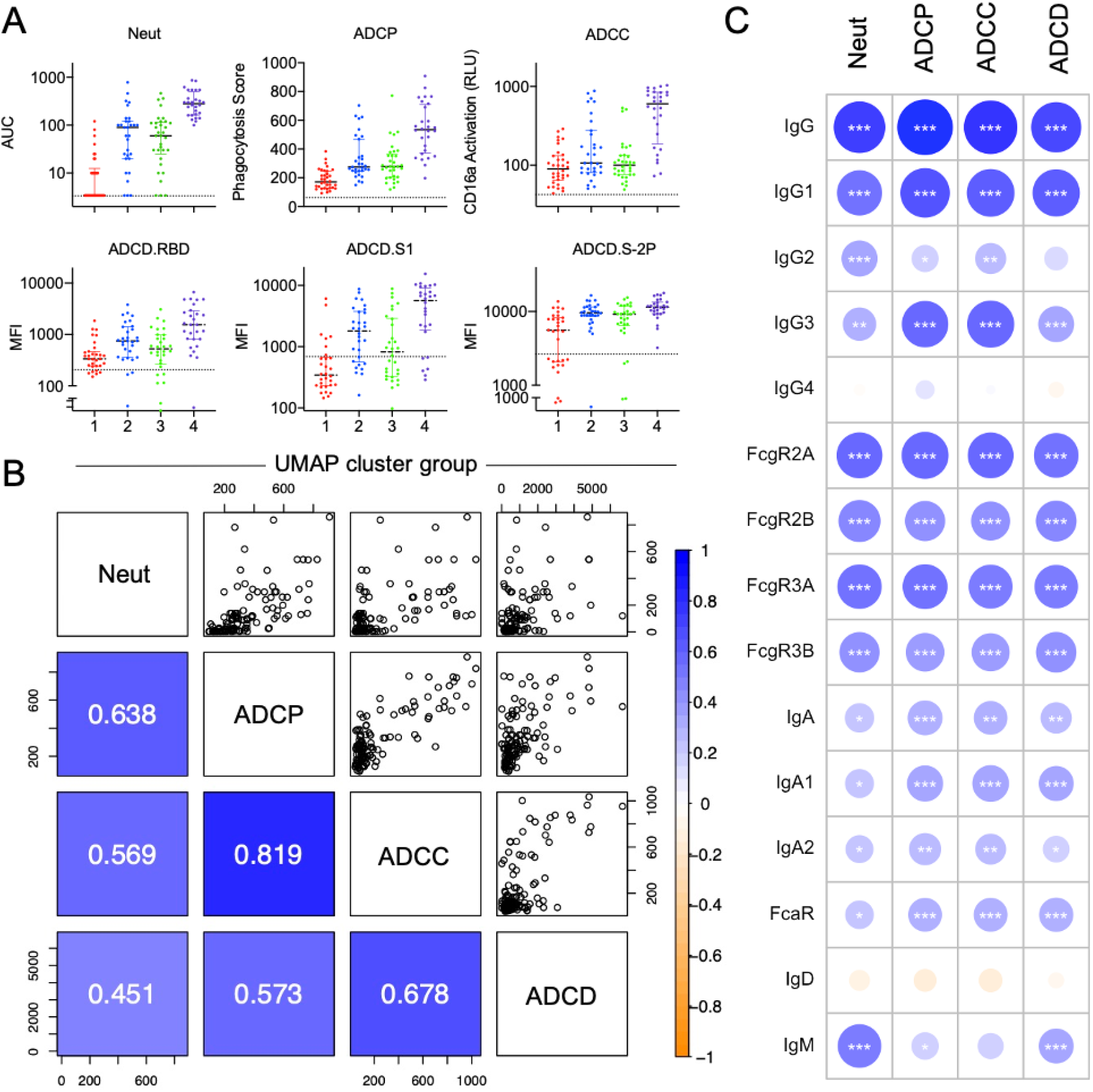
Functional Characterization of Plasma Antibodies. **A**. Neutralization, antibody-dependent cell-mediated phagocytosis (ADCP), antibody-dependent cellular cytotoxicity (ADCC), and antibody-dependent complement deposition (ADCD) activity of convalescent plasma donor samples, with donors colored by UMAP/k-means clusters. Dotted line indicates mean activity observed among naïve donor samples. **B**. Correlations between RBD-specific Ab features to functions in plasma, colored and labeled by Pearson correlation coefficient (R_P_) in the lower-left quadrant. **C**. Correlations (R_P_) between RBD-specific Fc array features and neutralization and effector functions. Significance of Pearson correlations (*p < 0.05; **p < 0.01; ***p < 0.001) are provided along with circles that are colored and sized according to their Pearson correlation coefficients (R_P_).

Because a number of the effector functions were tested specifically against the RBD antigen, we measured the degree and direction of correlation between RBD-specific Ab biophysical features and other Ab functions (**Figure 5C**). ADCP and ADCC were most strongly correlated with Fc*γ*R-binding antibodies, IgG1, and IgG3, which strongly ligate Fc*γ*R. Among FcR, correlations with activating Fc*γ*RIIa and Fc*γ*RIIIa were most strongly correlated, consistent with their known mechanistic relevance to ADCP and ADCC.

As previously observed in the DHMC cohort^29^, IgM positively correlated with neutralization activity. Relationships between serum IgA responses and antibody functions were considerably weaker than those with IgG responses. Correlative relationships with ADCD tended to be weaker, consistent with the strong dependence of this function on spatial aspects of avid antibody binding and immune complex formation that are typically captured by detection with the C1q, an initiator of the complement cascade that was not evaluated in this study.

### Multivariate modelling methods to predict functional responses

With the dual goals of better understanding the humoral response features that may drive complex antibody functions and enabling robust predictions from surrogate measures, we applied supervised machine learning methods to the JHMI cohort dataset and sought to evaluate generalizability of models predicting these activities by employing the DHMC cohort for validation. A regularized generalized linear modeling approach trained to utilize Fc Array features to predict each antibody function with minimal mean squared error was selected based on prior success in identifying interpretable factors that contribute to functional activity while avoiding overfitting^40^. A five-fold cross-validation strategy was applied, and comparison to models trained on permuted functional data established model robustness (**Figure 6A**). The cross-validated models trained on diverse data subsets showed consistent accuracy (measured by mean squared error) and good generalization when applied to the validation cohort (DHMC).

**Figure 6.**
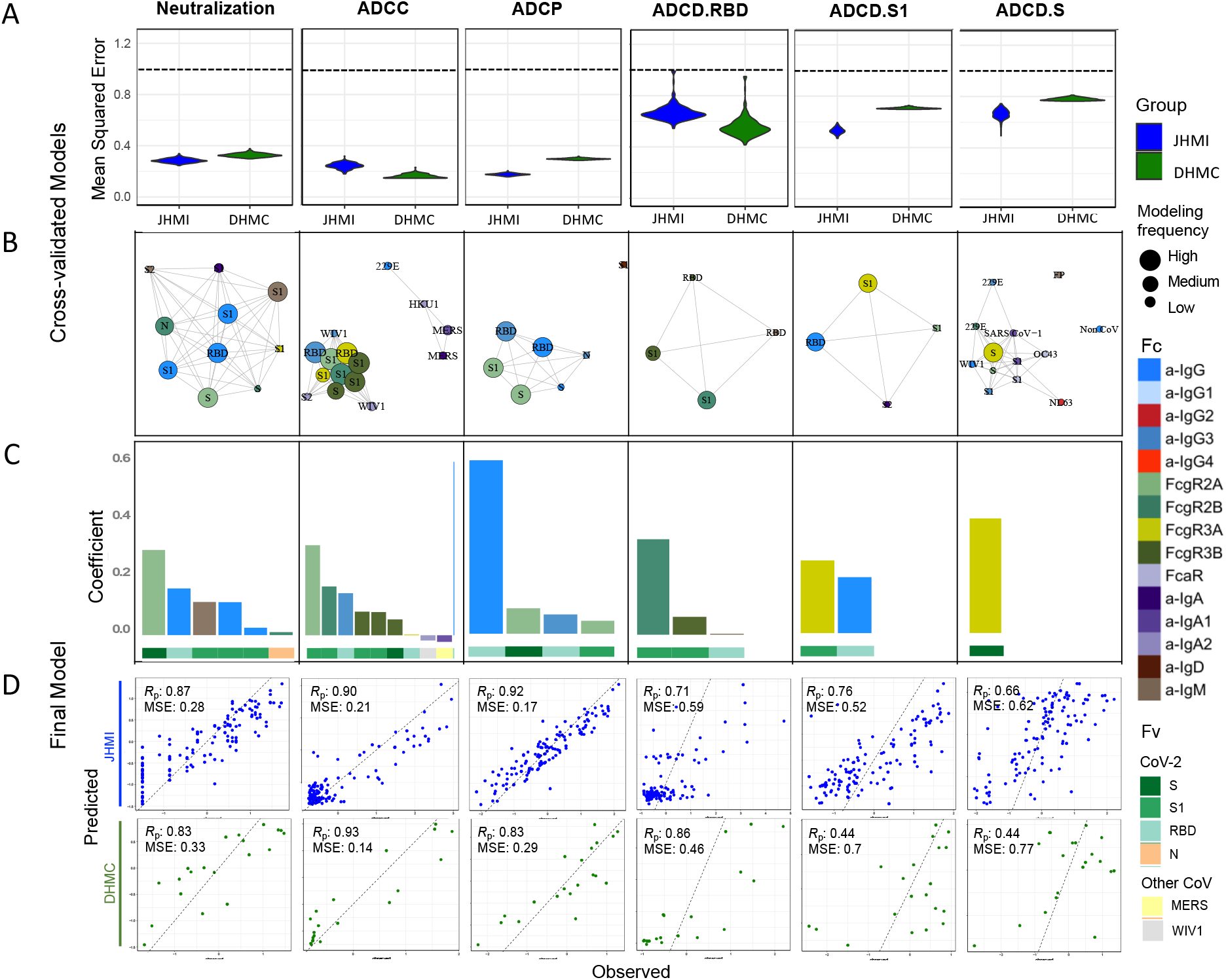
Multinomial linear regression modeling to predict functions using biophysical features. **A**. Comparison of mean squared error between testing (JHMI) and validation (DHMC) data sets for each functional assay. Dotted line indicates median performance on permuted data in the setting of repeated cross-validation. **B**. Network showing the identity, relative degree of correlation, and frequency with which features contribute to models in the setting of repeated cross-validation. **C**. Contribution of biophysical features to final models predictive of each function. **D**. Correlation between predicted and observed responses in the discovery (JHMI) and validation (DHMC) cohorts. Pearson correlation (R_P_) and means squared error (MSE) are reported in inset. Antigen specificity (Fv) and Fc characteristics (Fc) shown in color bars.

Weighted correlation network analysis (**Figure 6B**) demonstrated that a subset of features was consistently selected. The features that appeared with high frequency in repeated modeling were likely to have relatively high coefficients, and inversely, biophysical features with relatively small coefficients were prone to be influenced by the selected sample subset and to be removed by chance across the replicates. Collectively, frequently contributing features were exclusively related to spike recognition and were primarily driven by IgG and Fc*γ*R-binding antibodies. With this modeling approach, we were able to robustly evaluate the association between each functional assay and the selected biophysical features and gain insight into the relationships between antibody characteristics and functional activity.

Given established robustness, a final model for each function was trained on the complete JHMI cohort. Despite their sparseness compared to the control Ag, endemic CoV, and other epidemic CoV features, these models relied almost exclusively on antibody responses to the SARS-CoV-2 spike (**Figure 6C**). Consistent with our experimental approach, ADCC and ADCP models depended principally on antibodies specific to S1 and RBD. In contrast, the lead feature for virus neutralization was recognition of stabilized spike (S-2P). Similarly, complement deposition against whole spike was best predicted by a single feature related to spike trimer recognition. Responses to the S2 domain were not observed to contribute to functional predictions.

Beyond specificity, distinct antibody Fc characteristics contributed to model predictions. The most frequent Fc characteristic of features contributing to the final model of neutralization potency was the magnitude of IgG response, consistent with neutralization being FcR-independent. In contrast, the most frequent Fc characteristics in modeling ADCC and ADCP were Fc*γ*RIII- and Fc*γ*RII-binding responses, respectively, the receptors most relevant to each function. Further, despite comprising a relatively small fraction of circulating IgG, but consistent with its enhanced ability to drive ADCP^41^, IgG3 antibodies specific to RBD made a substantial contribution to models of ADCP activity, suggesting the potential importance of this particular subclass in the effector function of convalescent donor plasma. The link between neutralization and S1-specific IgM, an isotype typically associated with initial exposures^42^, suggests the possibility that these putative *de novo* lineages may exhibit superior neutralization activity and conversely that recalled, cross-reactive antibodies may be less likely to be neutralizing. However, a direct mechanistic contribution of IgM to neutralization potency cannot be excluded. Further studies are needed to investigate these alternative possibilities.

Lastly, correlation coefficients between the observed values of functional outcomes and the predicted results from the multivariate model were calculated, allowing for better visualization of model performance (**Figure 6D**). While functions were predicted with differing degrees of accuracy, all models generalized well to the independent validation cohort, and all relied upon features with established biological relevance. Reliable prediction of diverse antiviral activities from antibody profiles could contribute to donor prioritization strategies aimed at maximizing the global functionality of transfused plasma.

## Discussion

Convalescent plasma is one of the leading treatments of hospitalized patients for COVID-19. Following transfusion of more than 100,000 individuals in the United States with convalescent plasma, the FDA issued an Expanded Use Authorization. The largest multicenter study of over 35,000 patients suggested that early transfusion together with high titer units were needed for optimal clinical effect^11^. This observation estalblished a dose-response effect suggesting the existence of specific, measurable qualities that could be used to select the most effective plasma, but did not define a specific mechanism of action.

Relevant to convalescent plasma therapy and resistance of convalescent donors to reinfection, SARS-CoV-2-specific antibodies can elicit diverse antiviral functions beyond neutralization. These less well characterized functions were measured and related to biophysical antibody profiles. Multinomial linear regression identified distinct biophysical features that predicted antibody functions such as ADCC, ADCP, ADCD, and neutralization. Although models considered responses toward both endemic CoV and SARS-CoV-2, only SARS-CoV-2-specific responses were predictive of functional activity in independent discovery and validation cohorts. The consistency between antibody features contributing to each modeled function and expected biological relevance suggests that modeling approaches such as that employed here can identify mechanisms of antibody activity. Effector functions were most strongly correlated with Fc*γ*R-binding antibodies, IgG1, and IgG3. Neutralization was correlated with IgM responses, which may suggest the development of novel responses, as opposed to reactivation of responses to endemic CoV. SARS-CoV-2 specific IgM has also attracted interest because of its association with lower risk of death from COVID-19^9^. Non-neutralizing mechanisms of antibody-mediated protection against SARS-Cov-2 have not been extensively studied, but there is some evidence that both ADCC and phagocytosis can contribute antiviral effects against other coronaviruses^43–45^. Collectively, these functions have been suggested to play an important role in antibody-mediated defense against SARS-CoV-2 ^46^ and associated with vaccine-mediated protection^47,48^.

The antibody responses measured in convalescent subjects in this study were highly diverse, both in the SARS-CoV-2 antigens recognized and the magnitude of the responses; the latter observation is largely characteristic of the humoral responses measured to date ^9,10^. Interestingly, the magnitude of the responses against the spike protein of the endemic OC43, HKU1, and 229E were elevated relative to that of naïve subjects, suggesting that SARS-CoV-2 infection may boost a pre-existing population of cross-reactive B cells that target conserved CoV epitopes. The rapid rise in IgG by day 10-12 of infection^49^ rather than a response whereby IgM preceeds IgG is also consistent with an amnestic response. These observations suggest the “original antigenic sin” phenomenon, wherein antibody responses against earlier, related pathogens restrict the B cell repertoire available against novel infections, leading to boosting of those pre-existing antibodies at the expense of *de novo* antibody responses. However, by targeting conserved domains, these responses are typically not neutralizing since the receptor-binding domain tends to be highly variable. In some cases, this repurposing of the existing repertoire leads to a less effective antibody response against a new CoV^26^. Indeed, correlations between responses to SARS-CoV-2 antigens and endemic CoV suggest that recalled antibodies were more likely to recognize the S2 domain, and less likely to recognize the RBD, which is the target of most neutralizing antibodies isolated to date.

Limitations of this study range from cohort composition to the experimental and analytical approaches employed. Individuals in the naïve control cohort were generally younger and sourced from a different geographic location, which may impact our observation of apparent boosting of responses toward endemic CoV. Additionally, the convalescent and naïve subjects enrolled in the DHMC cohort provided serum samples, whereas the convalescent subjects in the JHMI cohort contributed plasma, which could result in differences in antibody detection and functional activity. Nevertheless, the model trained on convalescent plasma samples was able to make accurate predictions on convalescent serum samples. Recombinant antigen and lab-adapted cell lines were employed for several of the functional assays, and the substitution of surrogate measurements such as FcγRIIIa activation was made in place of target cell death. Thus, *in vitro* function may differ substantially from the *in vivo* processes these assays are meant to mimic. Given high feature dimensionality and relatively fewer subjects, LASSO regularization was used to increase the quality of prediction. This approach simplified the resulting models and improved interpretability of the selected variables, but tends to eliminate features that are highly correlated to selected variables in the established model, which can result in a trade-off between model simplification and obscuring potential biological mechanisms.

In summary, this study establishes three Fc-dependent activities in convalescent plasma beyond viral neutralization that could have antiviral effects against SARS-CoV-2, namely ADCC, phagoycotis and complement activation. These activities could explain therapeutic effects of plasma with low neutralizing capacity^11^. With this information we provide a proof of principle for the modeling of diverse antiviral activities against SARS-CoV-2 using biophysical inputs more amenable to high-throughput measurement. This work begins to define the specificities and Fc domain characteristics of antibodies associated with potent neutralization and effector function. However, therapeutically desirable plasma antibody functions have yet to be determined in humans. While a strong evidence base exists for the role of neutralizing antibodies in protection based on animal models and in the setting of human immune responses against other CoV^23,50–52^ and is beginning to accrue for SARS-CoV-2 infection^18,19,53^, continued analysis of the associations between passively transferred plasma characteristics and patient outcomes will likely be key to identifying the recipients who are most likely to benefit and the donors most likely to provide that benefit in the context of the COVID-19 pandemic.

## Methods

### Human subjects

The principal cohort of the study that was used for the training of the model has been previously described ^8^. Briefly, it comprised 126 adult subjects (mean age - 43 years; range - 19-78 years) previously diagnosed with SARS-CoV-2 infection by PCR+ nasal swab who met the standard eligibility criteria for blood donation and were collected in the Baltimore, MD and Washington DC area (Johns Hopkins Medical Institutions, JHMI cohort). The cohort was composed of 68 males (54.0%) and 58 females (46.0%). Eleven cases (8.7%) were severe enough to require hospitalization (mean duration of stay - 4 days; range 1-8 days). The cohort used for validation of the model comprised 20 SARS-CoV-2 convalescent individuals from the Hanover, New Hampshire area (Dartmouth Hitchcock Medical Center, DHMC cohort) (mean age - 40 years; range - 18-77); comprised 10 males and 10 females; among which 4 subjects (20%) were hospitalized. Infection with SARS-CoV-2 was confirmed in all convalescent subjects by real-time reverse-transcriptase–polymerase-chain-reaction of a nasopharyngeal swab. Plasma (JHMI) or serum (DHMC) was collected from each donor approximately one month after symptom onset or first positive PCR test in the case of mild or asymptomatic disease (**Supplemental Table 1**).

Human subject research was approved by both the Johns Hopkins University School of Medicine’s Institutional Review Board and the Dartmouth-Hitchcock Medical Center Committee for the Protection of Human Subjects. All participants provided informed written consent.

### Antigen and Fc Receptor expression and purification

Prefusion-stabilized, trimer-forming spike protomers (S-2P) of SARS-CoV-2; closely related and/or epidemic strains (SARS-CoV-1, WIV1, and MERS^54^); endemic CoV (229E, OC43, NL63, and HKU1); and a fusion of the receptor-binding domain of SARS-CoV-2 N-terminally to a monomeric human IgG4 Fc domain were transiently expressed in either Expi 293 or Freestyle 293-F cells, and purified via affinity chromatography, all according to the manufacturers’ protocols, as previously described ^29^. Human Fc*γ*R were expressed and purified as described previously^55^.

### Fc array assay

Coronavirus antigens – including S trimers, S subdomains (*i*.*e*., S1 and S2), and other viral proteins from SARS-CoV-2, plus the S trimers and subdomains from SARS CoV-1, MERS, HKU1, OC43, NL63, 229E, and WIV1 (**Supplemental Table 2**) – and the control antigens influenza HA and herpes simplex virus (HSV) gE proteins were covalently coupled to Luminex Magplex magnetic microspheres using a two-step carbodiimide chemistry as previously described^56^. Biotinylated SARS-CoV-2 fusion peptide was immobilized on neutravidin-coupled microspheres. Pooled polyclonal serum IgG (IVIG), the SARS CoV-1-specific monoclonal Ab CR3022 that cross-reacts with SARS-CoV-2 S ^57^, and VRC01, an HIV-specific monoclonal Ab, were used as controls to define bead antigenicity profiles. Pilot experiments were used to determine the optimal dilution of plasma for titrations. Test concentrations for plasma ranged from 1:250 to 1:5000 and were varied per detection reagent (**Supplemental Table 3**). Isotypes and subclasses of antigen-specific Abs were detected using R-phycoerthrin (PE) conjugated secondary Abs and by FcRs tetramers as previously described^55^. A FlexMap 3D array reader detected the beads and measured PE fluorescence in order to calculate the Median Fluorescence Intensity (MFI).

### Neutralization assay

Plasma from SARS-CoV-2 convalescent donors were tested in microneutralization assays using SARS-CoV-2/WA-1/2020 virus^8,58^ obtained from BEI Resources. VeroE6-TMPRSS2 cells were used to propagate the virus and to determined infectious virus titers using a 50% tissue culture infectious dose (TCID50) assay as previously described for SARS-CoV^8,58^ using Institutional Biosafety Committee approved protocols in Biosafety Level 3 containment. Two-fold dilutions of plasma were incubated with 100 50% tissue culture infectious units (TCID50s) for one hour at room temperature in a volume of 100 μL. The virus-plasma solution was then added to one well of VeroE6TMPRSS2 cells in a 96 well plate, incubated for 6 hours before being replaced with media. After incubation at 37°C for two days, the cells were fixed with 150 μL 4% formaldehyde followed by staining with Naptho blue black (Sigma Aldrich) and scoring for wells protected from infection. The assay was performed in hextuplicate and the area under the curve was calculated from the neutralizing antibody curve. Neutralization of the serum samples were tested using a VSV-SARS-CoV pseudovirus system as previously described^29,59^, and neutralization expressed as IC^60^ values.

### Phagocytosis assay

An assay of Ab-dependent phagocytosis by monocytes (ADCP) was performed essentially as described^60,61^. Briefly, 1 µm yellow-green fluorescent microspheres (Thermo, F8813) covalently conjugated with recombinant RBD were incubated for 3 hrs with dilute plasma specimens and the human monocytic THP-1 cell line (ATCC, TIB-202). After pelleting, washing, and fixing, phagocytic scores were calculated as the product of the percentage of cells that phagocytosed one or more fluorescent beads and the median fluorescent intensity of this population as measured by flow cytometry with a MACSQuant Analyzer (Miltenyi Biotec). CR3022 and VRC01 were used as positive and negative controls, respectively. Antibody-independent phagocytosis was measured from wells containing cells and beads, but no antibody.

### CD16 reporter assay

A Jurkat Lucia NFAT reporter cell line (Invivogen, jktl-nfat-cd16) was used to measure the ADCC potential, represented by the extent of FcγRIIIa activation, of each sample. Engagement of the cell surface receptor leads to the secretion of luciferase into the cell culture supernatant. The cells were cultured according to the manufacturer’s recommendations. One day prior to performing the assay, a high binding 96 well plate was coated with 1 µg/mL SARS-CoV-2 RBD and incubated at 4°C overnight. Plates were then washed with PBS + 0.1% Tween20 and blocked at room temperature for 1 hr with PBS + 2.5% BSA. After washing, dilute plasma and 100,000 cells/well in growth medium lacking antibiotics (with a total volume of 200 µL) were cultured at 37°C for 24 hrs. The following day, 25 µL of supernatant was drawn from each well and transferred to an opaque, white 96 well plate and 75 µL of QuantiLuc substrate was added. Luminescence was read immediately on a SpectraMax Paradigm plate reader (Molecular Devices) using 1 s of integration time. The reported values are the mean of three kinetic reads taken at 0, 2.5, and 5 min. Negative control wells substituted assay medium for antibody sample while cell stimulation cocktail (Thermo, 00-4970-93) plus an additional 2 μg/mL ionomycin were used to induce expression of the luciferase transgene as a positive control.

### Complement deposition assay

Antibody-dependent complement deposition (ADCD) was quantified essentially as previously described^62^. In brief, plasmas were heat-inactivated at 56°C for 30 min prior to a 2 hr incubation with multiplex assay microspheres at room temperature. After washing, each sample was incubated for 1 hour at room temperature with human complement serum (Sigma S1764) at a concentration of 1:50. Samples were washed, sonicated, and incubated for 1 hour at room temp with murine anti-C3b (Cedarlane CL7636AP) followed by anti-mouse IgG1-PE secondary Ab (Southern Biotech 1070-09) at room temp for 30 min. After a final wash and sonication, samples were resuspended in Luminex Sheath Fluid and complement deposition in the form of the median fluorescent intensity of the PE measured on a MAGPIX (Luminex Corp) instrument. Wells lacking Ab and but still containing heat-inactivated human complement serum served as negative controls.

### Data analysis and visualization

Basic analysis and visualization were performed using GraphPad Prism. Heatmaps, correlation plots, and boxplots were generated in R (supported by R packages pheatmap, corrplot, and ggplot2). Hierarchical clustering was used to cluster and visualize data using the Manhattan and Euclidean metrics. Fc Array features were filtered by elimination of features for which the samples exhibited signal within 10 standard deviations (SD) of the technical blank. Fc Array features were log transformed then scaled and centered by their standard deviation from the mean (z-score). A student’s two-tailed t-test with Welch’s correction with a cutoff of p=0.05 was used to define features different between groups. Pearson correlation coefficients were calculated for the correlation matrices.

UMAP was employed in the R package “umap” version 0.2.6.0 to enable dimensionality reduction of the JHMI Fc Array dataset. Upon log transformation, default UMAP parameters were used with the following exceptions: random_state = 45, min_dist = 1E-9, knn_repeats: -1, set_op_mix_ratio= 1. k-means was tested with a range of k = 1:15 to identify an optimal number of clusters as defined by a visual identification of an “elbow” in a plot of variance versus number of clusters. To identify features associated with each cluster, individual clusters were compared to the other three clusters using a student’s two-tailed t-test with Welch’s and Bonferroni’s corrections and a cutoff of p=0.05.

Multivariate linear regression was employed to predict the functional outcomes with the biophysical features, where an L1-penalization (LASSO) was applied to eliminate the variables that were less relevant to the outcome and reduce overfitting ^63^. By imposing a penalty on the absolute value of the feature coefficient, LASSO regression reinforces performance generalizability through its use of regularization and variable selection. The biophysical features from the JHMI and DHMC cohorts were log^10^ transformed to compensate for the positive skewness among the subjects in the Fc array dataset; functional measurements of ADCP, neutralization, and S1-specific ADCD were log^10^ transformed to reduce the prediction error of the models. All humoral responses in the two cohorts were centered and scaled independently. The lambda parameter (λ) was tuned using five-fold cross-validation with the biophysical features and functional measurements from the JHMI cohort. The progress of this refinement process was evaluated based on the mean squared error. A process of 200-times repeated modeling was used to investigate the potential of the different combinations of the biophysical features for modeling. Established with the JHMI cohort, a final model was selected based on the minimum MSE obtained among the repeated validations run on the DHMC cohort. The selected features and their coefficients were reported at “lambda.1se” to optimize the generalizability and provide more regularization to the model. Analysis was conducted with the R package “Glmnet”. The correlation network was conducted with the biophysical features that were repeatedly selected within the repeated modeling process. The “igraph” package was employed to calculate the weighted square adjacency matrix and create the network visualization.

## Data Availability

Data and code to reproduce analyses are available at (link pending).

## Data and Code Availability

Data and code to reproduce analyses are available at (link pending).

## Acknowledgements

We would like to thank all participants who enrolled in this study. VSV psuedovirus expression plasmids were provided by Dr. Michael Letko (Rocky Mountain Laboratories), CoV S-2P and RBD-Fc expression constructs were provided by Dr. Jason McLellan (UT Austin), and fusion peptide was provided by Dr. Laura Walker and Mrunal Sakharkar (Adimab). The following reagent was produced under HHSN272201400008C and obtained through BEI Resources, NIAID, NIH: Spike Glycoprotein Receptor Binding Domain (RBD) from SARS-Related Coronavirus 2, Wuhan-Hu-1 with C-Terminal Histidine Tag, Recombinant from Baculovirus, NR-52307. The following reagent was deposited by the Centers for Disease Control and Prevention and obtained through BEI Resources, NIAID, NIH: SARS-Related Coronavirus 2, Isolate USA-WA1/2020, NR-52281 This work was supported in part by the Division of Intramural Research, National Institute of Allergy and Infectious Diseases, as well as extramural support from the National Institute of Allergy and Infectious Diseases (R01AI120938, R01AI120938S1 and R01AI128779 to A.A.R.T, NIH Center of Excellence in Influenza Research and Surveillance HHSN272201400007C to A.P. and T32AI102623 to E.U.P.), National Heart Lung and Blood Institute (K23HL151826 to E.M.B), National Cancer Institute (2 P30 CA 023108-41 to M.E.A.), National Institute of General Medical Sciences (P20-GM113132 BioMT Molecular Tools Core) Bloomberg Philanthropies (A.C.) and Department of Defense (W911QY2090012 to A.C. and D.S.). A.C. was supported in part by NIH grants AI052733, AI15207 and HL059842. S.E.B. is supported by NIH NIAID 2T32AI007363.

## Author Contributions

Contributed samples – E.M.B., A.A.R.T., D.S., S.S.

Collected experimental data – H.N., A.R.C., S.E.B., R.I.C., W.W.-A., K.L, A.P

Performed data analysis – S.X., J.A.W.

Drafted the manuscript - H.N., A.R.C., S.E.B, S.X., J.A.W

Reviewed and edited the manuscript – all authors

Supervised research – M.E.A., P.F.W.

Conceived of work – M.E.A., A.D.R., A.A.R.T., A.C., H.B.L.

## Competing Interests Statement

The authors declare no competing interests.

## Notes

### Competing Interest Statement

The authors have declared no competing interest.

### Author Declarations

Human subject research was approved by both the Johns Hopkins University School of Medicine Institutional Review Board and the Dartmouth-Hitchcock Medical Center Committee for the Protection of Human Subjects. All participants provided informed written consent.

